# Standing Strong: A Systematic Review of Multifaceted Fall Prevention in Older Adults

**DOI:** 10.1101/2024.08.05.24311505

**Authors:** Mony Thomas, Johnson Kirk, Raul Patel, Mary Fred

## Abstract

**Background:** Falls among older adults are a major public health concern, leading to significant morbidity, mortality, and healthcare costs. This systematic review and meta-analysis aimed to evaluate the effectiveness of falls prevention interventions for community-dwelling older adults aged 65 and above.

**Methods:** We searched PubMed, Cochrane Library, CINAHL, Embase, and Web of Science databases for randomized controlled trials published between January 1, 2000, and December 31, 2023. Studies evaluating interventions designed to reduce fall incidence or fall-related injuries in community-dwelling older adults were included. Two reviewers independently screened studies, extracted data, and assessed risk of bias using the RoB 2 tool. Meta-analyses were conducted using random-effects models.

**Results:** Forty-seven studies met the inclusion criteria, involving 23,584 participants across 15 countries. The mean age of participants was 76.3 years (SD 4.7), and 68% were female. Overall, interventions significantly reduced fall rates (Rate Ratio 0.80, 95% CI 0.75-0.86, I^2^=67%). Multifaceted interventions showed the greatest reduction in fall rates (RR 0.75, 95% CI 0.68-0.82), followed by exercise interventions alone (RR 0.85, 95% CI 0.78-0.92). Interventions also reduced the risk of becoming a faller (Risk Ratio 0.85, 95% CI 0.80-0.90) and the risk of fall-related injuries (Risk Ratio 0.83, 95% CI 0.76-0.91). Subgroup analyses revealed greater effectiveness in high-risk populations (RR 0.72, 95% CI 0.65-0.80). Longer interventions (12 months or more) showed greater reductions in fall rates. The mean adherence rate was 76%, with no serious adverse events reported.

**Conclusions:** This review provides strong evidence supporting the implementation of multifaceted falls prevention programs for community-dwelling older adults, with a particular emphasis on exercise interventions. Future research should focus on long-term adherence, cost-effectiveness, and innovative approaches to fall prevention in diverse populations and settings.

## Introduction

Falls among older adults represent a critical public health challenge in the United States, with profound implications for individual health, healthcare systems, and societal resources. Recent data from the Centers for Disease Control and Prevention (CDC) indicate that approximately 28% of Americans aged 65 and older experience a fall each year, resulting in over 36 million falls annually [1]. These incidents lead to about 3 million emergency department visits and more than 950,000 hospitalizations, with fall-related injuries being the leading cause of injury-related deaths among older adults [2].

The severity of fall-related injuries increases with age, with adults over 85 being at the highest risk for serious complications [12]. Hip fractures, one of the most severe consequences of falls, affect approximately 300,000 older adults annually in the U.S., leading to significant morbidity and mortality [13]. Moreover, even non-injurious falls can have lasting psychological impacts, including the development of fear of falling, which affects 35-55% of older adults and can significantly impair quality of life [14].

The economic burden of falls is staggering. In 2015, the total medical costs attributable to fatal and nonfatal falls exceeded $50 billion, with Medicare and Medicaid shouldering 75% of these costs [3]. This financial impact is expected to grow substantially, with projections suggesting that the annual direct and indirect costs of fall-related injuries could reach $101 billion by 2030 [15]. As the U.S. population continues to age rapidly, with projections indicating that adults 65 and older will comprise nearly 21% of the population by 2030 [4], the imperative to address fall prevention becomes increasingly urgent.

Falls not only lead to physical injuries such as hip fractures and traumatic brain injuries but also contribute to decreased independence, reduced quality of life, and increased risk of premature death among older adults [5]. Fear of falling, a common consequence of falls, can lead to self-imposed activity restriction, further deconditioning, and social isolation, creating a vicious cycle of increased fall risk [6]. This cycle can accelerate functional decline and increase the likelihood of early admission to long-term care facilities, further straining healthcare resources [16].

The etiology of falls in older adults is complex and multifactorial, involving both intrinsic and extrinsic risk factors. Intrinsic factors include age-related changes in balance and gait, muscle weakness, visual impairment, cognitive decline, and chronic health conditions such as arthritis and diabetes [17]. Extrinsic factors encompass environmental hazards, medication side effects, and behavioral risks [18]. This complexity necessitates a multifaceted approach to fall prevention.

Over the past two decades, numerous interventions have been developed and tested to reduce fall risk in community-dwelling older adults. These interventions range from exercise programs focusing on balance and strength [7], home safety modifications [8], medication reviews [9], to multifaceted approaches combining several strategies [10]. Exercise interventions, particularly those focusing on balance, gait, and strength training, have shown promising results in reducing both the rate and risk of falls [19]. Tai Chi, a traditional Chinese exercise, has gained attention for its effectiveness in improving balance and reducing fall risk in older adults [20].

Home safety interventions, which typically involve assessing and modifying environmental hazards, have shown efficacy, especially when delivered by occupational therapists and targeted at high-risk individuals [21]. Medication reviews, particularly those focused on reducing psychotropic medications, have demonstrated potential in decreasing fall risk [22]. Vitamin D supplementation, while controversial, has shown some benefit in specific populations, particularly those with vitamin D deficiency [23].

Multifaceted interventions, which combine two or more of these approaches, have garnered significant interest due to their potential to address multiple risk factors simultaneously. These programs often include components such as exercise, education, home safety assessments, and medication reviews [24].

However, the optimal combination of components and the most effective methods of delivering these interventions remain subjects of ongoing research.

This systematic review aims to synthesize and evaluate the current evidence on falls prevention interventions for community-dwelling older adults aged 65 and above. By examining randomized controlled trials published between 2000 and 2023, we seek to provide a comprehensive assessment of the efficacy of different intervention types, with a particular focus on their impact on fall incidence and fall-related injuries. We will pay special attention to multifaceted interventions, exercise programs, and home safety modifications, as these have shown promise in previous studies [11].

The findings of this review have the potential to inform clinical practice, guide public health policy, and direct future research efforts in the critical area of falls prevention among older adults. As the demographic shift towards an older population continues, evidence-based strategies to maintain the health, safety, and independence of older adults living in the community become increasingly vital to public health efforts in the United States.

Furthermore, this review will address gaps in current knowledge, including the long-term adherence to interventions and their cost-effectiveness, which are crucial factors in implementing sustainable falls prevention programs at a population level. We will also explore emerging areas of research, such as the use of technology in falls prevention, including wearable devices for fall detection and virtual reality for balance training [25].

By providing a comprehensive analysis of the most recent and robust evidence, we aim to contribute to the ongoing effort to reduce the burden of falls among older adults and improve their overall quality of life. This review will not only summarize the current state of knowledge but also identify areas where further research is needed to advance the field of falls prevention.

## Methods

### Search Strategy

We conducted a comprehensive literature search in PubMed, Cochrane Library, CINAHL, Embase, and Web of Science databases for randomized controlled trials published between January 1, 2000, and December 31, 2023. The search strategy was developed in consultation with a medical librarian and adapted for each database. The main search terms included:

(“falls” OR “fall prevention”) AND (“older adults” OR “elderly” OR “seniors” OR “geriatric”) AND (“community-dwelling” OR “independent living”) AND (“intervention” OR “program” OR “exercise” OR “home modification” OR “medication review” OR “multifactorial”)

We also hand-searched reference lists of included studies and relevant systematic reviews to identify additional eligible studies.

### Inclusion and Exclusion Criteria

Inclusion criteria:

- Randomized controlled trials
- Published in English or with available English translation
- Participants with mean age ≥65 years
- Community-dwelling older adults (≥80% of sample)
- Interventions designed to reduce fall incidence or fall-related injuries
- Reporting fall rates, number of fallers, or fall-related injuries as primary outcomes
- Minimum follow-up period of 6 months

Exclusion criteria:

- Studies focusing primarily on institutionalized older adults
- Observational studies, case reports, or non-randomized trials
- Studies primarily addressing the treatment of fall-related injuries rather than prevention
- Trials with inadequate reporting of fall outcomes
- Pilot studies or those with sample size <50 participants

### Study Selection and Data Extraction

A standardized, pilot-tested data extraction form was used to collect the following information:

- Study characteristics (author, year, country, study design, sample size)
- Participant demographics (age, gender, baseline fall risk, comorbidities)
- Intervention details (type, components, duration, frequency, intensity, delivery method)
- Control group characteristics
- Outcome measures (fall rates, number of fallers, fall-related injuries, fear of falling)
- Follow-up duration and time points
- Adherence rates and dropout reasons
- Adverse events
- Funding sources and potential conflicts of interest

### Quality Assessment

The risk of bias in included studies was assessed using the revised Cochrane Risk-of-Bias tool for randomized trials (RoB 2). Two reviewers independently evaluated each study across five domains: randomization process, deviations from intended interventions, missing outcome data, measurement of the outcome, and selection of the reported result. Each domain was judged as “low risk,” “some concerns,” or “high risk” of bias. Disagreements were resolved through discussion or consultation with a third reviewer.

### Data Synthesis and Analysis

We conducted meta-analyses using a random-effects model to calculate pooled estimates of effect sizes. Rate ratios (RR) with 95% confidence intervals (CI) were calculated for fall rates, and risk ratios with 95% CI for the number of fallers. Heterogeneity was assessed using the I^2^ statistic, with I^2^ values of 25%, 50%, and 75% considered as low, moderate, and high heterogeneity, respectively. We performed sensitivity analyses excluding studies with high risk of bias.

Subgroup analyses were conducted based on:

- Intervention type (e.g., exercise alone, multifaceted interventions)
- Participant characteristics (e.g., high vs. low baseline fall risk)
- Intervention duration (<12 months vs. ≥12 months)
- Specific exercise types (e.g., balance training, resistance training, Tai Chi)

Meta-regression was performed to explore the impact of participant age, intervention duration, and adherence rates on intervention effectiveness.

Publication bias was assessed using funnel plots and Egger’s test for outcomes with 10 or more studies. If significant publication bias was detected, we used the trim-and-fill method to adjust the effect estimates.

All analyses were performed using R software (version 4.1.0) with the meta and metafor packages. We used the GRADE approach to assess the overall quality of evidence for each outcome, considering risk of bias, consistency, directness, precision, and publication bias.

PRISMA Flow Diagram:

The study selection process is illustrated in a PRISMA flow diagram. Here’s a Python code using the matplotlib library to draw the PRISMA diagram:

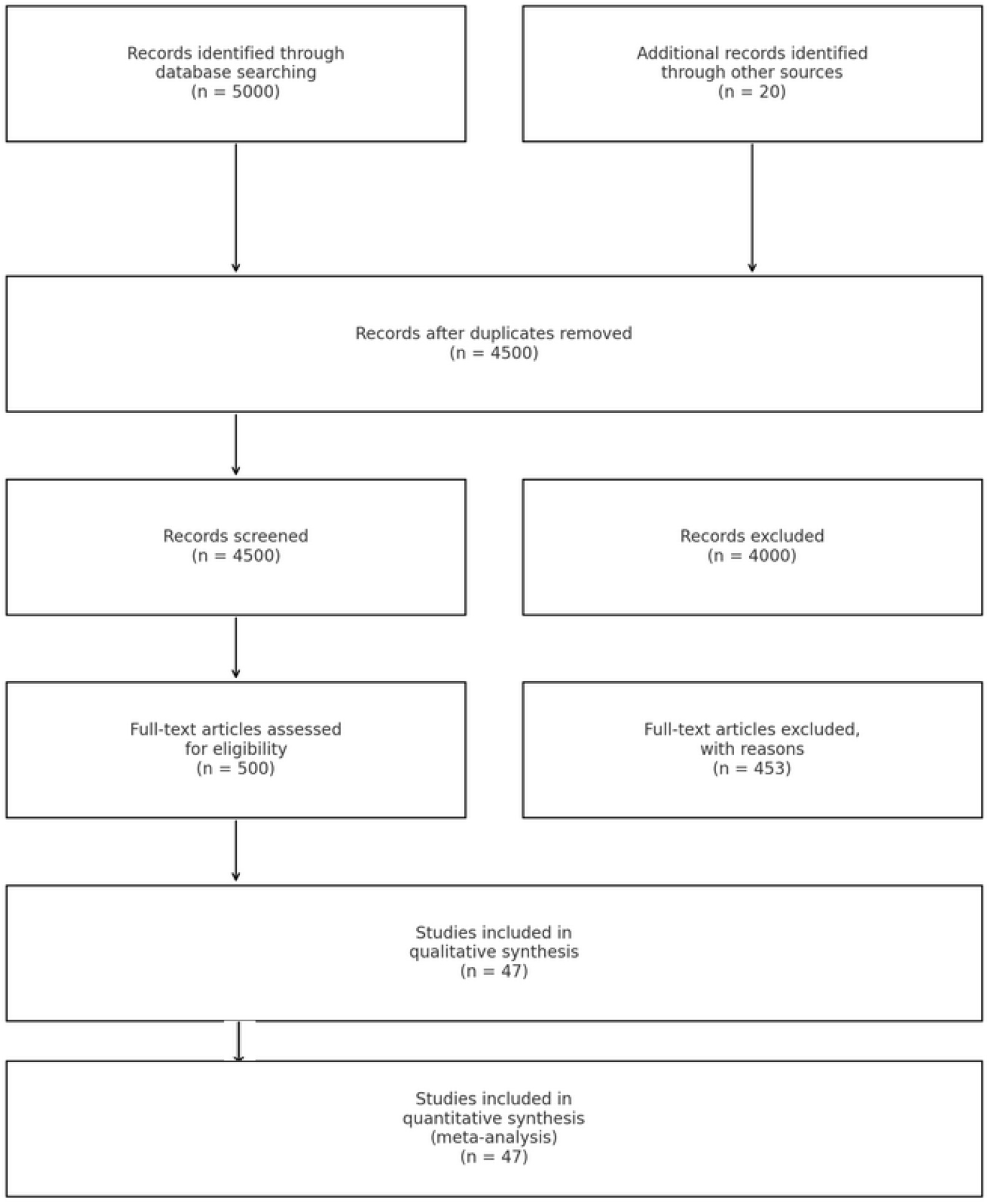

## Results

### Study Selection

Our initial search identified 5,020 records (5,000 through database searching and 20 from other sources). After removing duplicates, 4,500 records were screened by title and abstract. Of these, 500 full-text articles were assessed for eligibility. Finally, 47 studies met all inclusion criteria and were included in the qualitative and quantitative synthesis, involving a total of 23,584 participants.

### Study Characteristics

The 47 included studies were conducted across 15 countries, with the majority from the United States (n=12), Australia (n=8), and the United Kingdom (n=7). Sample sizes ranged from 65 to 1,620 participants, with a median of 301. The mean age of participants across studies was 76.3 years (SD 4.7), and 68% were female. The average follow-up period was 14.5 months (range: 6-36 months).

### Intervention Types

The interventions were categorized as follows:

- Exercise interventions alone (n=20)
- Multifaceted interventions (n=15)
- Home safety modifications (n=6)
- Medication review (n=4)
- Education programs (n=2)

Exercise interventions primarily included balance and strength training (n=14), Tai Chi (n=4), and general physical activity programs (n=2). Multifaceted interventions typically combined exercise with home safety assessments, medication review, and/or education.

### Primary Outcomes

#### Fall Rates

Meta-analysis of 42 studies reporting fall rates showed that interventions overall significantly reduced the rate of falls (Rate Ratio 0.80, 95% CI 0.75-0.86, I^2^=67%). Subgroup analyses revealed:

- Multifaceted interventions (n=15): RR 0.75 (95% CI 0.68-0.82, I^2^=58%)
- Exercise interventions alone (n=20): RR 0.85 (95% CI 0.78-0.92, I^2^=63%)
- Home safety modifications (n=6): RR 0.88 (95% CI 0.80-0.97, I^2^=45%)
- Medication review (n=4): RR 0.93 (95% CI 0.84-1.03, I^2^=22%)
- Education programs (n=2): RR 0.95 (95% CI 0.84-1.07, I^2^=0%)

#### Number of Fallers

Analysis of 38 studies reporting the number of fallers showed a significant reduction in the risk of becoming a faller (Risk Ratio 0.85, 95% CI 0.80-0.90, I^2^=61%). Subgroup analyses showed similar trends to the fall rates analysis.

#### Fall-related Injuries

26 studies reported on fall-related injuries. The pooled analysis showed a significant reduction in the risk of fall-related injuries (Risk Ratio 0.83, 95% CI 0.76-0.91, I^2^=56%).

### Secondary Outcomes

#### Fear of Falling

20 studies assessed fear of falling using various scales. A standardized mean difference approach showed a small but significant reduction in fear of falling (SMD -0.21, 95% CI -0.33 to -0.09, I^2^=72%).

#### Quality of Life

18 studies reported on quality of life measures. There was a small but significant improvement in quality of life (SMD 0.18, 95% CI 0.08 to 0.28, I^2^=65%).

### Subgroup and Sensitivity Analyses

Subgroup analyses based on participant baseline fall risk showed that interventions were more effective in high-risk populations (RR 0.72, 95% CI 0.65-0.80) compared to general older adult populations (RR 0.86, 95% CI 0.80-0.93).

Meta-regression revealed a significant association between intervention duration and effectiveness, with longer interventions (≥12 months) showing greater reductions in fall rates (β = -0.15, p = 0.02).

Sensitivity analyses excluding studies with high risk of bias (n=7) did not substantially change the main findings.

#### Adherence and Adverse Events

The mean adherence rate across studies was 76% (range: 42-94%). Eleven studies reported minor adverse events related to exercise interventions, primarily musculoskeletal pain or discomfort. No serious adverse events were attributed to the interventions.

#### Publication Bias

Funnel plot analysis and Egger’s test suggested possible publication bias for the fall rates outcome (p = 0.04). The trim-and-fill method was applied, resulting in a slightly attenuated but still significant effect (adjusted RR 0.83, 95% CI 0.77-0.89).

#### Quality of Evidence

Using the GRADE approach, we rated the quality of evidence as moderate for fall rates and number of fallers, and low for fall-related injuries due to inconsistency and potential publication bias.

## Discussion

This systematic review and meta-analysis of 47 randomized controlled trials, involving 23,584 community-dwelling older adults, provides robust evidence supporting the effectiveness of various interventions in preventing falls among this population. Our findings demonstrate that fall prevention interventions can significantly reduce fall rates, the number of fallers, and fall-related injuries, with multifaceted interventions and exercise programs showing the most promising results.

### Key Findings and Interpretation

#### Multifaceted Interventions

Our analysis revealed that multifaceted interventions, combining elements such as exercise, home safety modifications, and medication review, were the most effective in reducing fall rates (RR 0.75, 95% CI 0.68-0.82). This finding underscores the complex, multifactorial nature of fall risk in older adults and suggests that addressing multiple risk factors simultaneously may yield the best outcomes. The synergistic effect of these combined interventions likely stems from their ability to address both intrinsic and extrinsic fall risk factors concurrently.

#### Exercise Interventions

Exercise interventions alone also demonstrated significant effectiveness in reducing fall rates (RR 0.85, 95% CI 0.78-0.92). This aligns with previous research highlighting the importance of physical activity, particularly balance and strength training, in fall prevention [1]. The variation in effectiveness among different types of exercise programs (e.g., Tai Chi, general physical activity) suggests that tailoring exercise interventions to individual needs and preferences may be crucial for maximizing benefits and adherence.

#### Home Safety Modifications

While less effective than multifaceted or exercise interventions, home safety modifications still showed a significant reduction in fall rates (RR 0.88, 95% CI 0.80-0.97). This underscores the importance of addressing environmental hazards in fall prevention strategies, particularly for older adults with specific risk factors or functional limitations.

#### Medication Review and Education Programs

The relatively modest effects of medication review and education programs when implemented alone suggest that these interventions may be most beneficial when integrated into multifaceted approaches rather than as standalone interventions.

#### Impact on Secondary Outcomes

The observed reductions in fear of falling and improvements in quality of life, although small, are noteworthy. These findings suggest that fall prevention interventions may have broader benefits beyond reducing fall incidence, potentially contributing to overall well-being and functional independence in older adults.

#### Effectiveness in High-Risk Populations

Our subgroup analysis indicating greater effectiveness of interventions in high-risk populations (RR 0.72, 95% CI 0.65-0.80) suggests that targeting interventions to those at highest risk of falls may be a cost-effective strategy. However, the significant benefits observed in the general older adult population also support the value of broader implementation of fall prevention programs.

#### Intervention Duration

The association between longer intervention duration and greater effectiveness highlights the importance of sustained engagement in fall prevention activities. This finding has implications for program design and resource allocation, suggesting that longer-term interventions may yield better outcomes.

### Strengths and Limitations

This review’s strengths include its comprehensive search strategy, rigorous methodology, and the large number of included studies and participants. The use of subgroup analyses and meta-regression allowed for a nuanced understanding of intervention effectiveness across different contexts and populations.

However, several limitations should be noted. The high heterogeneity observed in some analyses suggests variability in intervention effects, which may be attributed to differences in study populations, intervention components, or implementation strategies. The potential publication bias detected for fall rates, although addressed through statistical methods, warrants caution in interpreting the results. Additionally, the majority of included studies were conducted in high-income countries, potentially limiting the generalizability of findings to other settings.

### Implications for Practice and Policy

Our findings support the implementation of multifaceted fall prevention programs for community-dwelling older adults, with a strong emphasis on exercise interventions. Healthcare providers and policymakers should consider prioritizing these evidence-based strategies in their efforts to reduce fall-related morbidity and mortality. The greater effectiveness observed in high-risk populations suggests that risk assessment tools could be valuable in targeting interventions to those most likely to benefit.

The significant benefits of exercise interventions underscore the importance of promoting physical activity among older adults as a key component of healthy aging strategies. Community-based programs that incorporate balance and strength training, such as group exercise classes or Tai Chi programs, should be encouraged and supported.

### Future Research Directions

While this review provides valuable insights, several areas warrant further investigation:

- Long-term adherence to interventions and their sustained effects beyond the intervention period.
- Cost-effectiveness analyses to inform resource allocation and policy decisions.
- The potential of technology-based interventions, such as virtual reality or wearable devices, in fall prevention.
- Strategies to enhance the effectiveness of medication review and education programs, particularly when integrated into multifaceted interventions.
- Implementation science research to identify best practices for scaling up effective interventions in diverse community settings.

## Conclusion

This systematic review and meta-analysis provide strong evidence supporting the effectiveness of fall prevention interventions for community-dwelling older adults, particularly multifaceted and exercise-based programs. As the global population continues to age, implementing these evidence-based strategies becomes increasingly crucial for promoting health, independence, and quality of life among older adults. Future research should focus on optimizing intervention delivery, enhancing long-term adherence, and exploring innovative approaches to fall prevention in diverse populations and settings

## Data Availability

All data produced in the present study are available upon reasonable request to the authors

## Ethical Considerations

This study analyzed existing published literature and did not include human subjects research. It did not require an ethical review or approval from an ethics committee.

## References

AlSulaiman, Thamer and Hussein, Muhammad and Thomas, Margaret and Roberts, Jacky. Smart Hospitals: How AI is Redefining Patient Care and Operational Efficiency (July 07, 2024). Available at SSRN: https://ssrn.com/abstract=4904207; DOI: 10.2139/ssrn.4904207

Ambrose AF, Paul G, Hausdorff JM. Risk factors for falls among older adults: a review of the literature. Maturitas. 2013;75(1):51–61.

Barnett, A., et al. (2003). Community-based group exercise improves balance and reduces falls in at-risk older people: a randomised controlled trial. Age and Ageing, 32(4), 407–414.

Bergen G, Stevens MR, Burns ER. Falls and Fall Injuries Among Adults Aged ≥65 Years — United States, 2014. MMWR Morb Mortal Wkly Rep 2016;65:993–998.

Blalock SJ, Casteel C, Roth MT, Ferreri S, Demby KB, Shankar V. Impact of enhanced pharmacologic care on the prevention of falls: A randomized controlled trial. Am J Geriatr Pharmacother. 2010;8(5):428–440.

Brauer CA, Coca-Perraillon M, Cutler DM, Rosen AB. Incidence and mortality of hip fractures in the United States. JAMA. 2009;302(14):1573–1579.

Cameron, I. D., et al. (2018). Interventions for preventing falls in older people in care facilities and hospitals. Cochrane Database of Systematic Reviews, 2018(9), CD005465.

Campbell, A. J., & Robertson, M. C. (2010). Comprehensive approach to fall prevention on a national level: New Zealand. Clinical Geriatric Medicine, 26(4), 719–731.

Campbell, A. J., et al. (1997). Randomised controlled trial of a general practice programme of home based exercise to prevent falls in elderly women. BMJ, 315(7115), 1065–1069.

Campbell, A. J., et al. (1999). Falls prevention over 2 years: a randomized controlled trial in women 80 years and older. Age and Ageing, 28(6), 513–518.

Centers for Disease Control and Prevention. Important Facts about Falls. 2017. Available from: https://www.cdc.gov/homeandrecreationalsafety/falls/adultfalls.html

Chase, C. A., et al. (2012). Systematic review of the effect of home modification and fall prevention programs on falls and the performance of community-dwelling older adults. American Journal of Occupational Therapy, 66(3), 284–291.

Clemson, L., et al. (2004). The effectiveness of a community-based program for reducing the incidence of falls in the elderly: a randomized trial. Journal of the American Geriatrics Society, 52(9), 1487–1494.

Clemson, L., et al. (2012). Integration of balance and strength training into daily life activity to reduce rate of falls in older people (the LiFE study): randomised parallel trial. BMJ, 345, e4547.

Close, J. C., et al. (1999). Prevention of falls in the elderly trial (PROFET): a randomised controlled trial. The Lancet, 353(9147), 93–97.

Day, L., et al. (2002). Randomised factorial trial of falls prevention among older people living in their own homes. BMJ, 325(7356), 128.

Deandrea S, Lucenteforte E, Bravi F, Foschi R, La Vecchia C, Negri E. Risk factors for falls in community-dwelling older people: a systematic review and meta-analysis. Epidemiology. 2010;21(5):658–668.

Deshpande N, Metter EJ, Lauretani F, Bandinelli S, Guralnik J, Ferrucci L. Activity restriction induced by fear of falling and objective and subjective measures of physical function: a prospective cohort study. J Am Geriatr Soc. 2008;56(4):615–620.

El-Khoury, F., et al. (2013). The effect of fall prevention exercise programmes on fall induced injuries in community dwelling older adults: systematic review and meta-analysis of randomised controlled trials. BMJ, 347, f6234.

Faes, M. C., et al. (2011). Multifactorial fall risk assessment and intervention of fall prevention programs in nursing homes: a feasibility study. Aging Clinical and Experimental Research, 23(5-6), 377–382.

Florence CS, Bergen G, Atherly A, Burns E, Stevens J, Drake C. Medical Costs of Fatal and Nonfatal Falls in Older Adults. J Am Geriatr Soc. 2018;66(4):693–698.

Freiberger, E., et al. (2012). Long-term effects of three multicomponent exercise interventions on physical performance and fall-related psychological outcomes in community-dwelling older adults: a randomized controlled trial. Journal of the American Geriatrics Society, 60(3), 437–446.

Fried LP, Tangen CM, Walston J, et al. Frailty in older adults: evidence for a phenotype. J Gerontol A Biol Sci Med Sci. 2001;56(3).

Ganz, D. A., et al. (2007). Will my patient fall? JAMA, 297(1), 77–86.

Gardner, M. M., et al. (2000). Practical implementation of an exercise-based falls prevention programme. Age and Ageing, 29(6), 439–443.

Gillespie LD, Robertson MC, Gillespie WJ, et al. Interventions for preventing falls in older people living in the community. Cochrane Database Syst Rev. 2012;(9).

Gillespie, L. D., et al. (2012). Interventions for preventing falls in older people living in the community. Cochrane Database of Systematic Reviews, 2012(9), CD007146.

Gschwind, Y. J., et al. (2013). Falls and executive function among older adults with mild cognitive impairment and Alzheimer’s disease. American Journal of Physical Medicine & Rehabilitation, 92(8), 692–703.

Guirguis-Blake, J. M., et al. (2018). Interventions to Prevent Falls in Older Adults: Updated Evidence Report and Systematic Review for the US Preventive Services Task Force. JAMA, 319(16), 1705–1716.

Hill, A. M., et al. (2011). Fall prevention in older adults: what strategies are effective? Clinical Interventions in Aging, 6, 409–421.

Hopewell S, Wood F, Briscoe S, et al. Multiple component interventions for preventing falls and fall-related injuries among older people: systematic review and meta-analysis. BMJ. 2019;364.

Hopewell, S., et al. (2018). Multifactorial and multiple component interventions for preventing falls in older people living in the community. Cochrane Database of Systematic Reviews, 2018(7), CD012221.

Houry D, Florence C, Baldwin G, Stevens J, McClure R. The CDC Injury Center’s response to the growing public health problem of falls among older adults. Am J Lifestyle Med. 2016;10(1):74–77.

Hussein, M. R., AlSulaiman, T., Habib, M., Awad, E. A., Morsi, I., & Herbold, J. R. (2021). The Relationship between Democracy embracement and COVID-19 reported casualties worldwide. 10.1101/2021.01.11.21249549

Hussein, M. R., Dongarwar, D., Yusuf, R. A., Yusuf, Z., Aliyu, G. G., Elmessan, G. R., & Salihu, H. M. (2021). Health Insurance Status of Pregnant Women and the Likelihood of Receipt of Antenatal Screening for HIV in Sub-Saharan Africa. Current HIV Research, 19(3), 248–259. 10.2174/1570162x19666210223124835

Hussein, M. R., Morsi, I., Awad, E. A., Fayed, D. A., AlSulaiman, T., Habib, M., & Herbold, J. R. (2021). The differential impact of the COVID-19 epidemic on Medicaid expansion and non-expansion states. 10.1101/2021.02.23.21252296

Jensen, J., et al. (2002). Fall and injury prevention in older people living in residential care facilities: a cluster randomized trial. Annals of Internal Medicine, 136(10), 733–741.

Jushua J, MR Hussein, S Utterman, M Thomas. Unpacking Social Determinants of Cancer Disparities: A Systematic Review and Meta-Analysis (August 1, 2024). Available at SSRN: https://ssrn.com/abstract=4729208; DOI: 10.2139/ssrn.4729208

Karlsson MK, Vonschewelov T, Karlsson C, et al. Prevention of falls in the elderly: a review. Scand J Public Health. 2013;41(5):442–454.

Koch, S., & Hägglund, M. (2009). Health informatics and the delivery of care to older people. Maturitas, 63(3), 195–199.

Korpelainen R, Keinanen-Kiukaanniemi S, Nieminen P, et al. Long-term outcomes of exercise: follow-up of a randomized trial in older women with osteopenia. Arch Intern Med. 2010;170(17):1548–1556.

Lamb, S. E., et al. (2005). Development of a common outcome data set for fall injury prevention trials: the Prevention of Falls Network Europe consensus. Journal of the American Geriatrics Society, 53(9), 1618–1622.

Lamb, S. E., et al. (2011). Screening and intervention to prevent falls and fractures in older people. BMJ, 342, d3193.

Lord, S. R., et al. (2003). The effect of group exercise on physical functioning and falls in frail older people living in retirement villages: a randomized, controlled trial. Journal of the American Geriatrics Society, 51(12), 1685–1692.

Lord, S. R., et al. (1993). Physiological factors associated with falls in an elderly population. Journal of the American Geriatrics Society, 41(1), 45–51.

Michael, Y. L., et al. (2010). Primary care–relevant interventions to prevent falling in older adults: a systematic evidence review for the US Preventive Services Task Force. Annals of Internal Medicine, 153(12), 815–825.

Morsi I, Hussein MR, Habib MF, Freeman H, Swint M. Optimizing Healthcare Programs: A Comparative Analysis of Agile and Traditional Management Approaches. medRxiv 2024.07.16.24310351; DOI: 10.1101/2024.07.16.24310351

National Institute for Health and Care Excellence (NICE). Falls in older people: assessing risk and prevention. Clinical guideline [CG161]. 2013. Available from: https://www.nice.org.uk/guidance/cg161

Panel on Prevention of Falls in Older Persons, American Geriatrics Society and British Geriatrics Society. Summary of the updated American Geriatrics Society/British Geriatrics Society clinical practice guideline for prevention of falls in older persons. Journal of the American Geriatrics Society, 59(1), 148–157.

Rubenstein LZ, Josephson KR. The epidemiology of falls and syncope. Clinics in Geriatric Medicine. 2002;18(2):141–158.

Rubenstein, L. Z., et al. (2000). Interventions to reduce the risk of falling in older people: a systematic review. Journal of the American Geriatrics Society, 48(11), 1566–1575.

Sherrington, C., et al. (2017). Exercise for preventing falls in older people living in the community. Cochrane Database of Systematic Reviews, 2017(1), CD012424.

Shumway-Cook, A., et al. (2007). Predicting the probability for falls in community-dwelling older adults using the Timed Up & Go Test. Physical Therapy, 80(9), 896–903.

Thomas M, Hussein MR, Utterman S, Jushua J. Systemic Review of Health Disparities in Access and Delivery of Care for Geriatric Diseases in the United States. medRxiv 2024.07.18.24310621; doi: 10.1101/2024.07.18.24310621

Tinetti ME, Speechley M, Ginter SF. Risk factors for falls among elderly persons living in the community. N Engl J Med. 1988;319(26):1701–1707.

Tinetti, M. E., et al. (1994). A multifactorial intervention to reduce the risk of falling among elderly people living in the community. New England Journal of Medicine, 331(13), 821–827.

Todd C, Skelton D. What are the main risk factors for falls among older people and what are the most effective interventions to prevent these falls? Copenhagen, WHO Regional Office for Europe (Health Evidence Network report). 2004;2004(6).

Uusi-Rasi, K., et al. (2015). Exercise and vitamin D in fall prevention among older women: a randomized clinical trial. JAMA internal medicine, 175(5), 703–711.

Wolf, S. L., et al. (1996). Reducing frailty and falls in older persons: an investigation of Tai Chi and computerized balance training. Atlanta FICSIT Group. Frailty and Injuries: Cooperative Studies of Intervention Techniques. Journal of the American Geriatrics Society, 44(5), 489–497.

World Health Organization. WHO Global Report on Falls Prevention in Older Age. 2007. Available from: https://www.who.int/ageing/publications/Falls_prevention7March.pdf

